# “Move the ultrasound scanning machine to where the woman is”: Stakeholder perspectives and experiences of task-sharing antenatal ultrasound scanning with trained midwives in Blantyre, Malawi

**DOI:** 10.64898/2025.12.18.25342641

**Authors:** Linly Seyama, Chipiliro Payesa, Fidelis Sindani, Limbanazo Matandika, Blessings Kapumba, Lingstone Chiume, Yamikani Chimwaza, Luis Gadama, Pooja Sripad, Stephanie Suhowatsky, Lisa Noguchi, Sufia Dadabhai

## Abstract

In 2018, Malawi adopted the World Health Organization antenatal care (ANC) recommendation of at least one ultrasound scan (USS) for pregnant women before 24 weeks gestation. A shortage of trained providers and USS machines, however, has significantly hindered implementation. A qualitative study explored diverse stakeholder perspectives on integrating USS into ANC services through task-sharing with trained midwives in 10 primary health centers in Blantyre, Malawi. We conducted seven focus group discussions with 58 stakeholders, including midwives, current ANC clients, policymakers, male partners of pregnant women, women that had given birth in the last 16 weeks, nurse/midwife educators, and community representatives. Discussions were audio-recorded with consent, transcribed and translated. Topics included acceptability and feasibility of task-sharing, experience with point-of-care USS (POCUS) devices and recommendations for scale-up. Data were coded in NVivo and thematically analyzed. Four themes emerged. [1] Community sensitization and male involvement are needed for successful integration of antenatal USS and scale-up of task-sharing with midwives. [2] Stakeholders perceive that antenatal USS has a positive impact on continuity and quality of care, specifically early detection of complications, women’s birth preparedness, and referral-related cost- and time-savings. [3] Main barriers to USS implementation were unmet expectations among clients for USS results counselling, increased midwife workloads and client wait times, midwives perceptions about the challenging nature of the training content, and device-specific challenges. [4] Integrated scale-up requires the inclusion of USS in midwives’ official scope of practice, more POCUS devices, and standardized training of all midwives in Malawi to provide USS at the primary care level. Stakeholders supported task-sharing of USS to midwives and integration into routine ANC. They provided valuable feedback and specific recommendations for successful scale-up.

## Introduction

Antenatal care (ANC) services are essential for maternal health because they offer an opportunity for early identification of risk factors in pregnancy [1] and increase women’s chances of being assisted by a skilled healthcare provider in the event of abnormal findings [2]. The 2016 World Health Organization (WHO) ANC guidelines recommend, and at least one ultrasound scan (USS) before 24 weeks of gestation to estimate gestational age, improve detection of anomalies and multiple pregnancies, minimize post-term labor inductions, and enhance patient-centered care pregnancy care[3]. The Malawi Ministry of Health adopted the WHO guidelines in 2018 [1]. Implementation of the USS standard of care in ANC has not been possible in most facilities due to the lack of equipment and trained personnel.

In Malawi, 80% of ANC services are provided at primary care facilities (sometimes secondary) by nurses and midwives [3]. They are not trained to provide basic obstetric USS, and it is not part of their scope of practice. USS services are provided by a limited number of sonographers, radiographers, obstetricians and obstetrics and gynecology (Ob/Gyn) registrars who primarily work in tertiary hospitals. Task-sharing the provision of USS with midwives is a plausible way of ensuring that this service is provided on a larger scale to pregnant women in Malawi. Task-sharing has shown to be efficient and effective in healthcare service delivery in areas with inequitable healthcare worker distributions [1], particularly for HIV testing, HIV and tuberculosis management, and hypertensive disorders in pregnancy[2–4] There is growing evidence on task-sharing for basic obstetric USS in the sub-Saharan Africa region. The ARC-005 study hypothesized that training midwives coupled with the availability and affordability of POCUS devices could make task-sharing possible [1,2].

A mixed methods implementation science study was conducted among trained midwives who used the point of care ultrasound (POCUS) device to increase the availability of USS services in routine ANC. Jhpiego and the Johns Hopkins Research Project, with the support of the Reproductive Health Department in the Ministry of Health, conducted the study with funding from The Bill and Melinda Gates Foundation, through the Antenatal/Postnatal Research Collective (ARC). Quantitative results on feasibility and acceptability have been reported by Payesa et al [5]. The objective of this qualitative study was to describe stakeholders’ perspectives, attitudes and experiences with USS by midwives with a POCUS device during routine ANC at health centers. Stakeholders represented health system, community and beneficiary perspectives, and provided feedback on and strategies for scale-up throughout Malawi.

## Methods

### Study Design

This multi-phase, mixed method prospective implementation science study investigated the feasibility and acceptability of the limited introduction of USS with the POCUS Butterfly iQ device into routine ANC service delivery at ten health centers in Blantyre District, Malawi. The study had three phases. In this study, we report on Phase 3.

Phase 1 - Training: Training of midwives on basic obstetric scanning and related anatomy, POCUS device use, integration into ANC workflow, counselling pregnant women and partners on USS results and referral within network. Midwives attended a 5-day in-person theoretical training facilitated by eight senior Obstetrics and Gynecology residents from Queen Elizabeth Central Hospital.

Phase 2 - Iterative Service Delivery: 1500 basic obstetric scans delivered by 45 midwives over approximately 8 months with supportive supervision and service delivery adaption as needed.

Phase 3 - Qualitative Evaluation: Evaluating the potential integration of task-sharing USS to midwives into current policies and practices for training and service provision. We conducted seven focus group discussions (FGDs) with stakeholders involved in the first two phases: midwives, policymakers, trainers in nursing/midwifery institutions, and clients, including pregnant women and community members. The FGDs enabled us to collect perspectives, attitudes and experiences with task-sharing of USS by midwives, including integration into routine care and strategies for scale-up throughout Malawi.

### Study Setting and Study Population

The study was conducted in Blantyre District in the Southern Region of Malawi. We purposively sampled groups to represent either implementers and those responsible for service delivery design or community including those who benefit from scanning services. Specifically, we conducted two FGDs among the midwives trained to provide based obstetric USS during the study (n=2); one with policymakers (n=1); one with Ob/Gyn registrars (n=1); one with nursing school educators (n=1) one with currently pregnant women that had received an ultrasound either on study or through other programs (n=1); and one with community representatives (n=1).

Apart from two national-level policymakers and two persons representing nurse/midwife training institutions, all other FDG participants were recruited from within Blantyre District. Community Engagement Officers purposively selected participants for the FGDs based on their role in the study or in the community. Participants included Ob/gyn registrars from Queen Elizabeth Central Hospital (Malawi’s southern region’s tertiary referral hospital) that led midwives’ USS training in Phase 1; ANC clients who the midwives had scanned in Phase 2; national and district-level policymakers knowledgeable about the safe motherhood agenda in Malawi; pregnant women from the surrounding community that had never received a USS; husbands/male partners of currently-pregnant women; women who delivered in the past 16 weeks; representatives from Malawi’ midwifery and nursing schools; and community representatives such as clergy, village chiefs and other traditional leaders. Midwives that participated in Phases 1 and 2 were eligible for recruitment into the FGDs; they represented both rural and urban areas of the District, specifically the heath facilities in Limbe, Mpemba, Chavala, Chimembe, Zingwangwa, Dziwe, South Lunzu, Mdeka, Lundu, and Mbayani.

Each focus group had 6 to 10 participants. Sampling was sufficient to draw out collective community and beneficiary as well as implementing and oversight stakeholder perspectives of incorporating USS as a part of ANC.

### Data Collection

FGDs were conducted from 27 February 2023 to 21 April 2023 by experienced and trained moderators (SS, WP, and LS) using a semi-structured interview guide (supplemental information). SS is PhD student at Liverpool University; LS was the project lead for the study, with a masters’ degree; while WP has a bachelor’s degree. The three moderators had research ethics certifications, ample qualitative data collection experience, and proficiency in both English and Chichewa languages. They were trained on the study objectives and FGD guides. They had no pre – existing relationship with participants. Moderators were asked to pretest the FGD guides and provide feedback to the research team in order to refine the questions and probes, to determine the estimated duration of FGDs, and to ensure they understood the motivation for each question.

Face-to-face FGDs were conducted in quiet meeting rooms at Johns Hopkins Research Project administration office with adequate spacing, adequate acoustics for audio-recording and sufficient privacy and door signs to minimize disruptions. They were administered in English, Chichewa or a mixture based on participants’ choice. Refreshments were provided to participants along with the Malawi kwacha equivalent of 10 USD to cover participants’ transport and time. The policymakers FGD (led by moderator LS) was conducted virtually to accommodate the various geographic locations of the participants. FGD discussion guides covered the following domains:

- Factors supporting the feasibility of midwives to conduct scans, counsel and refer women when needed (and related barriers).
- How to plan for scale-up of ANC ultrasound to more health centers/more women.
- Features of the training program for midwives & scope of practice & mentoring.
- Positive and negative experiences for women getting scanned & overall effect on ANC experience.
- Community reactions to ANC ultrasound scanning.

To reduce burden on participants, the length of the FGDs was targeted for one hour. FGDs were audio recorded, and recordings were transcribed verbatim and translated into English by dedicated, experienced contractors fluent in both English and Chichewa. Transcripts went through quality control by study site staff and the moderators before they were coded and analyzed.

### Data Coding and Analysis

Thematic analysis of transcript data was undertaken in the following five phases: initial coding framework creation, coding, intercoder reliability and agreement assessment, coding framework modification and coding of the entire dataset [6,7], and final code categorization. Two researchers (BK and LM) developed the codebook by independently coding four transcripts. Line-by-line transcripts were coded, and data were organized into themes and sub-themes. We conducted thematic coding in NVivo 14 using broad inductively defined themes (such as views on continuity of care and acceptance of ultrasound scanning or perceived benefits of including ultrasound scanning in antenatal care) and inductively derived sub-themes (such as changes to ANC service provision or improvement on maternal and neonatal outcomes). To improve inter-coder reliability, the coding of these interview transcripts and emerging themes were discussed with the entire study team (BK, LM, LS). Data saturation was achieved once all themes emerging from the data had been adequately captured and no new themes emerged. Thematic analysis and interpretation resulted in the final themes presented in this study.

The coding was refined until an acceptable level of consensus and inter-coder reliability were achieved. As interview transcripts were coded, the codebook was expanded as required to ensure that novel responses were captured for analysis. Previously coded transcripts were checked to code any novel responses that had not been captured. After coding all the transcripts, the coded data in NVivo was merged, and codes were categorized into broader themes to address the research questions around stakeholder perspectives, attitudes and experience. These included community-orientated factors, perceived impact on quality of care, barriers to implementation, and implications for integration and scale up. We also used framework matrices to compare perspectives between different stakeholders. Research findings were presented to participants for validation via member-checking, and quotations were presented to illustrate the findings for credibility and to ensure that participants’ perspectives were equitably represented [6,7]

### Ethical Consideration

Study participation was voluntary, and written informed consent was obtained from all participants before the start of each FGD. Ethics approval was obtained from the College of Medicine Research Ethics Committee (P.11/20/3196) and the Johns Hopkins School of Public Health Institutional Review Board (15449/CR987).

## Results

Fifty-eight (58) participants were included in the study: 20 midwives across two FGDs, eight registrars, six policymakers, seven women who received a scan in Phase 2; six representatives of training institutions, and 11 community representatives including recently-delivered women, clergy, village chiefs and men whose partners were pregnant. Among the 58 participants, 67% were females, with a mean age of 35 years. FGDs lasted from 34 to 77 minutes.

Four themes emerged from our final thematic analysis: the importance of community sensitization in the scale-up of antenatal USS; the perceived impact of USS on continuity and quality of care; recommendations for improving USS implementation; and implications of task-shifting for USS integration into ANC refer to Figure 1.

**Figure 1:**
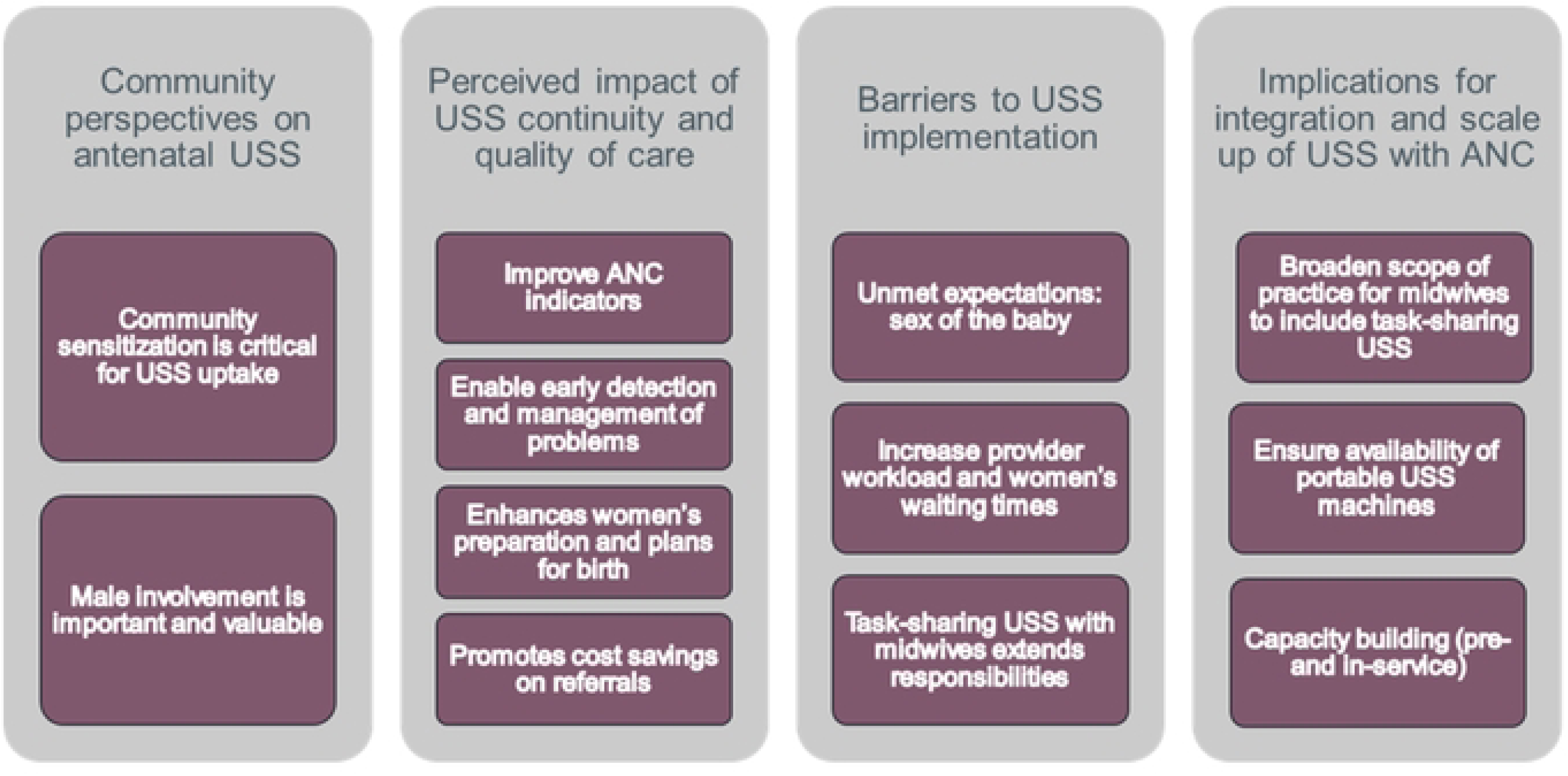
Themes and subthemes that emerged from focus group discussion data

### Theme 1: Community Perspectives Towards Antenatal Ultrasound Scanning

#### Community sensitization is critical for USS uptake

All participants were asked about their perceptions regarding USS and the rollout of USS services within health centers. Pregnant women, midwives, and community representatives indicated that community sensitization using various social modalities—social groups, village savings banks, or leaders—was critical for broad USS uptake. Most participants mentioned that community members need to know what USS is for, what information it can reveal to a pregnant woman and her baby, what it cannot show, and why the ANC visits may take longer.

> “I think that the other way of encouraging pregnant women to accept ultrasound scanning is through some of the groupings that women participate in; we have social groups, we have village savings banks, we have saving groups in communities, and sometimes we have time to sit down and chat after laundry. And those of us who are not religious leaders or chiefs, we have a role supporting those people that, the time that we are chatting, we could discuss with other women about ultrasound scanning, or maybe to those mothers that don’t know or to those that fail to understand properly what ultrasound scanning is.” (Community representative)

> “I am saying this because chiefs are influential in their communities, and community members tend to listen to the chiefs effectively, saying that once a chief has said something, people tend to do that thing without any delays at all. So, the chiefs can play a key role in helping to encourage the pregnant women to be going to access ultrasound scanning services.” (Pregnant woman).

#### Male involvement is important and valuable

Participants reported male involvement as important and valuable in USS because men help in decision-making regarding the uptake of health services and preparation for delivery. In this regard, they mentioned that it’s important to incorporate men into the messages about USS so that they, too, can become knowledgeable about the new development.

> “My thoughts are that if you go there with your husband, at the antenatal care clinic, your husband should also be allowed into the scanning room so that when you are being provided with the service the husband should be able to observe what is taking place. (Pregnant woman)

> Yes, as women we are indeed comfortable with having our husbands or partners present when accessing this service because if we access these services alone we find that we struggle to explain this service as well as the outcome we have had as a result of accessing this service to them when we get back home. (Pregnant woman)

When asked about strategies for raising awareness among men about obstetric USS, most participants indicated that deliberate efforts should be made to target men in their usual community gathering places including residential areas, funerals or church meetings. Others indicated that sharing information at the health facility when men have escorted their partners for ANC would help.

> “So, men are also available on Sunday in churches. Maybe they could make announcements in church saying that men should remain behind. And then, maybe a health worker should teach them, and in so doing it (information about antenatal USS) might spread faster.” (Community representative).

> “My view is that the good way is by calling men through the chiefs because it might not be possible for health workers to carry out door-to-door visits and find men at home, it’s not common for men to be at home”(Community representatives).

### Theme 2: Perceived impact of ultrasound scanning on continuity and quality of care

This section highlights changes that USS by midwives may have on the ANC quality and continuity of care. Participants reported several benefits from integration of USS on ANC indicators, early detection and management of problems, women’s preparation for birth and cost savings on referrals”

#### Improved ANC indicators

All participants reported that USS by midwives in health centers is a welcomed development in healthcare delivery systems. Midwives, policymakers, training institution representatives, and registrars agreed that moving USS closer to the point of care of pregnant women would improve ANC indicators, reduce complications, and improve maternal and neonatal outcomes.

> “So, it also helps that we should be able to detect early if there are some anomalies and that we should also be able to assist them early and avoid preventable maternal deaths” (USS-trained Midwife)

> “It’s a good idea that these midwives should be providing the ultrasound scanning so that we can at least improve this indicator of antenatal women attending antenatal care in the first trimester; it will boost this antenatal care indicator” (Training institution representative).

#### Enable early detection and management of problems

A key benefit of training midwives in USS was the early confirmation of pregnancies and the ability of midwives to provide more focused advice to women during their pregnancy. Both midwives and pregnant women noted that pregnancies were confirmed sooner than before USS when women had to wait until their pregnancy had progressed to be confirmed through palpation of a gravid uterus [typically later in the second trimester]. Participants further mentioned knowing the baby’s gestational age and size enabled early identification of complications and facilitated early referral if needed.

> “In the past when pregnant women came in during the first trimester of their pregnancies, we could not palpate the unborn child through a physical examination, and if we had no pregnancy test kits, we were just sending the women back telling them [to] come back after three or four months. However, with the coming of the Butterfly, we can provide the ultrasound scanning services there and then, and we can tell them outcomes…informing a pregnant client that [she] is indeed pregnant for this many months.” (Midwives)

> “…in the past, if a pregnant client had a complication, that complication was discovered maybe when it was already too late because the pregnancy had already grown. Now they can discover the complication before the pregnancy grows. In so doing they can find ways to start assisting you.”(Pregnant woman)

#### Enhances women’s preparation and plans for birth

Most participants noted that bringing USS closer to women not only enabled access but also aided them in preparing adequately for their upcoming baby and delivery. Continuing to provide Midwives also report that USS during routine ANC will allow women to plan their delivery plan early, such as transport and location.

> “There is no need to [for] the pregnant woman to go where the ultrasound scanning machine is. What we can do now is to move the ultrasound scanning machine to where the woman is.” (Midwives)

> “I think that ultra-sound scan should continue because it would help pregnant mothers to prepare accordingly, and maybe when that scan shows that there are twins, therefore the pregnant mother would prepare for twin babies.” (Midwives)

Midwives, pregnant women, and community members stated that training midwives to conduct USS benefits early ANC initiation and improved ANC attendance. Although the study saw few first-trimester pregnancies at the ANC, some participants mentioned that USS would help bring in first-trimester pregnant women for ANC services.

> Scanning is also helping to increase the turn-up of women, and especially when it comes to early antenatal care attendance, because most of the women start to attend antenatal care late. (Midwives)

#### Promotes cost savings on referrals

Many participants mentioned cost savings from referrals as a benefit, explaining that prior to the introduction of USS at the primary level, families incurred higher transport costs and time because pregnant women needed to travel to secondary or tertiary facilities to scan referrals.

> “It [training midwives in USS] will also help in reducing costs, men were in serious problems. And I also have some experience in that, because my wife had the same experience during her first pregnancy. She was transferred from [one] health facility coming here. In such a case it means that you have to travel from [name] district to visit your wife… if we know most of the things at the health center level, then it will be very helpful.” (Community)

Midwives and policymakers reported that scanning at primary-level facilities will reduce unnecessary referrals and congestion at secondary and tertiary facilities.

> “… we have reduced unnecessary referrals. Our facility is some 75 kilometers away from [name] Hospital. So, to just refer a client unnecessarily for that distance, it’s something challenging for those clients. And for them to prepare, the guardian, the children, the household and what have you”.(Midwives)

### Theme 3: Challenges with USS identified by midwives and pregnant women

Pregnant women who accessed scanning services, along with midwives, highlighted unmet expectations, prolonged waiting times, increased workloads, and difficulties with the devices as the primary challenges met.

#### Unmet expectations

Most midwives and pregnant women mentioned that many women requested the USS to determine the sex of the baby. Pregnant women wanted to know the sex of their unborn babies. However, this was not under the midwives’ scope of USS training or required service provision.

> “… but most of the clients would like to know about the sex of the baby when they come for antenatal care. When they come to the health facility, they would like to know about the sex of the baby, whether male or female, and they could come only for that reason…” (Pregnant women)

Community members indicated that they felt uncomfortable regarding the issue of scanning and counselling pregnant women about the sex of the unborn baby. They believed it might create conflicts in families that anticipate a specific gender of the child baby.

> “Most of the time when women are pregnant, they look for affection from the husband. So, sometimes a family could have female children only, and they may be thinking that their current pregnancy would give them their desires baby, but may be after scanning, the results show contrary to the expectation, this may discourage the couple and some men may ignore their pregnant partner because of the results of a scan.” (Community)

#### Increased provider workloads and women’s waiting time

Nearly all participants expressed dissatisfaction with the waiting time.

> “So, when there are fewer providers, it becomes somehow a challenge because ultrasound scans take some time, you spend more time with a client than the time that you would have spent if you were not providing scanning service” (Midwives).

> “I can say that now we are indeed being delayed” (Pregnant woman)

Some community members felt that with the introduction of the scanning device, women spent more time with midwives compared to previous check-ups. However, participants were also concerned about women spending more total time at the facility. Some participants [pregnant women and midwives] mentioned that the USS service extended waiting times. Conversely, other midwives suggested that there had been little change in the time spent with clients. They noted that the scan should not affect the timing of ANC visits, as this method was intended to replace hand palpation and essentially requires the same amount of time.”

> “So, at the moment it’s not different because, when they are offering ultrasound scan, they don’t use that gadget that they put in the ear and listen to. They just do a physical examination and do ultrasound scan, and then we come back, the difference is not so much, and it’s just a few minutes’ difference”. (community)

The increased workload for midwives was primarily noted by midwives, policymakers, and representatives from training institutions. Most of the participants pointed out the increased workload among midwives, which was mentioned regarding the time midwives spent with one client and at ANC

> “So, when there are fewer providers, it becomes somehow a challenge because ultrasound scans take some time, you spend more time with a client than the time that you would have spent if you were not providing scanning service.” (Midwives)

#### Task-sharing USS with midwives extends responsibilities

Some midwives, policymakers and training institution representatives indicated that while task-sharing of recommended USS as a part of comprehensive ANC services was possible, it added an additional responsibility for the ANC provider and increased their workload. Some registrars involved with the training suggested the content may be too advanced for midwives. Most participants, especially midwives, indicated that as much as the USS is a welcome service, the number of nurses providing it is inadequate because not all midwives were trained per facility. Adding USS responsibility to only a select few overwhelmed nurses at the primary level was challenging; however, participants concur that burden could be eased if more providers were trained.

> “We are saying that we are reducing the workload for the facilities that we used to refer these pregnant clients to. At the same time, we are increasing the workload in our facilities, which is why there is a need to train other service providers. So that there can be many healthcare providers that can provide this service.” (Midwives).

### Theme 4: Implications for integration and scale up of USS with ANC ultrasound at health centers

Registrars, policymakers, and representatives from training institutions were asked whether they believed it was necessary to enable the integration and scale up of USS in routine ANC, including expand midwives’ practice scope, necessary equipment supply, and nature and type of training required.

#### Broaden scope of practice for midwives

Policymakers, training institution representatives, and registrars stated that to scale up USS, the scope of practice must be re-defined, and competencies must be aligned for midwives.

Participants further mentioned that regulatory bodies such as the Nurses and Midwives Council and Medical Council will need to certify the proposed in-service training of midwives.

> “So, it’s not only in-service but also pre-service, because whatever the nurses had been trained in-service should align with their scope and competencies for midwives in Malawi… that should be clear” (Policy maker).

Registrars who conducted the USS training also noted that the content should be streamlined after narrowing the scope of practice for midwives. They believed that some of the material was too technical for the midwives.

> “I think there will be a need for some time to say, what content was delivered to our nurses, and what are the things that we need to know. And what are the things that they don’t need to know? Because, just as my colleagues have said, they were also trained in some complicated stuff.” (Registrars)

#### Ensure availability of portable USS devices

Midwives, registrars, and policymakers indicated that introducing portable and easy-to-use devices will facilitate the scaling up of ultrasound scan services. Providing ultrasound scan services in rural facilities requires portable devices because most of these facilities do not have adequate space for storage or to place traditional equipment device.

> “On the portability; for example, when traditional ultra-sound scans are placed in antenatal, then they cannot be moved to labor ward, for instance, if you would like to scan a pregnant mother.” (Midwives)

#### Capacity building [pre- and in-service]

Capacity building was reported as the main facilitator for integration and scale up. Almost all participants mentioned that it would be beneficial if all primary health facilities and midwives who provide ANC were trained to provide USS service.

> “It is a welcome development and if the midwives are well equipped to be scanning, especially at the primary level, it is going to be a very good development.” (Policymaker)

> As such, we have seen that even the midwives at the peripherals, were able to diagnose complications like breach presentations…[or]…placenta previa. Midwives at the very end, were able to diagnose and refer timely for further management. And it also helped in decongesting our tertiary facility.” (Policy maker)

Some participants believed incorporating the training into the nursing training curriculum is necessary to scale up USS provided by midwives.

> “It can be very beneficial if the issues to do with ultrasound scanning service provision were included in the syllabus, the syllabus used in the nursing schools. So when people are leaving these nursing schools, they should at least have some knowledge about the provision of this service. In doing so, once they get to the facilities they have been deployed to work in, they can just continue from what they already know since they already started learning about this service in the training schools, and not that they should start getting trained from scratch.” (Midwives)

When respondents were asked about personnel and training length, content, and target groups to improve USS provision in routine ANC, registrars, midwives, and representatives from training school institutions agreed that a few days of training were insufficient. Instead, they proposed that midwives partake in 1-2 weeks of training to adequately master scanning skills.

> “That being said, scaling up is a necessary thing that needs to be done, but for one, I felt like the training and practice period for the nurses was not enough, because I can tell you that, by the time we were finishing doing the visits, some of them were not confident to do the scanning. While we have some that, halfway through the process, you would know that this one would give you a proper scan.” (Registrars)

Midwives and representatives from training institutions emphasized the need to define trainers and mentors. They proposed that sonographers and practicing radiographers be designated as the primary trainers. Regarding mentors, no specific group was mentioned to serve as mentors apart from the registrars.

> “I think the best people to train these are the group we suggested, the registrars in obstetrics, the specialists in obstetrics. The sonographer, if it is a radiographer, should be someone who has been trained in this before he can go and say I have to train.

> Because, if you want to be a trainer you should not have the basic knowledge like the one you are training. You should have a little bit more knowledge and a better understanding of things. That is why we are specifying that specialists in obstetrics, registrars and sonographers, and radiographers should be trained as trainers, after being trained and not just direct”. (Training institution representative).

The cadre of nurses/midwives to be trained was also discussed, and most participants indicated that all nurses who provide ANC service should be trained. However, some participants mentioned that only some nurses - Nurse Midwife Technicians (NMTs) and registered nurses who provide service to pregnant women at the primary level - need to be trained in providing USS service.

> “I would suggest that maybe we have the NMTs and registered nurses because, at the primary level, we have more of the NMTs than the registered nurses, we have few at the primary level like my health center. So, I would suggest that we can start from nurse midwife technician, and then the registered nurses.” (Policy Maker)

## Discussion

Despite the demonstrated benefits of basic obstetric ultrasound (USS), uptake, adoption, scale-up, and compliance remain low in many low- and middle-income countries, including Malawi, where pregnancy complications are prevalent. Midwife-led USS is a potential strategy to increase access for pregnant women. This study explored the perspectives of key stakeholders on implementing and scaling up midwife-led basic obstetric USS in Blantyre, Malawi.

Our findings indicate that training midwives to provide obstetric USS is highly acceptable and potentially feasible for meeting the needs of pregnant women, particularly for one scan before 24 weeks of gestation. Community sensitization and male involvement are important for successful implementation of USS task-sharing with midwives. To scale up ANC ultrasound services to more facilities and patients, it is necessary to expand the scope of practice of midwives/nurses; develop focused, standardized training curricula; train more midwives; and plan for an infusion of ultrasound devices. Integration of USS into ANC was believed to improve quality of care, reduce complications through early detection and management. Registrars, sonographers, and experienced radiographers were recommended as trainers and mentors, consistent with ISUOG recommendations [8,9]. Both registered nurses and nurse midwife technicians could be trained.

Participants emphasized the role of community sensitization in order to ensure community awareness of any new service or product before initiation, to promote acceptance and uptake and to reduce misconceptions [8]. Findings suggest that community leaders have more influence in the uptake of new health services because people trust them even more than direct communication from health care workers. This aligns with the Sustainable Development Goals notion of integrated, people-centered health services, in which community participation is necessary and merits priority in achieving universal health coverage [2,9,10]. Our study identified strategies to involve men, who are critical in decision-making in the household, especially during pregnancy, including approaching men in clubs, churches, workplaces, and uniquely at funerals. In Malawi, funeral ceremonies are well-attended and often serve as platforms to share information that community members need to know. These findings are consistent with previous studies highlighting the importance of community involvement in the introduction of new health technologies and services, particularly in low-income settings [9].

USS task-sharing is acceptable and can enhance continuity of care and quality, improve ANC indicators, reduce complications, improve maternal and neonatal outcomes, facilitate early identification of complications and referral, confirm pregnancy, save money, and ensure easy access. These positive perceived impacts identified in our study are consistent with published data[1,11–14]. Additionally, our study found reduced congestion at the referral facility to be a notable benefit. The study revealed that before USS task-sharing, women were referred to secondary or tertiary facilities for obstetric USS. Some reasons for referral included confirming pregnancy or multiple gestations, which could be conducted at the primary facility if the service is available; concurrently, this was congesting the recipient facilities. We hypothesized that clients would receive timely attention in addition to decongesting referral facilities due to the reduced number of cases. This means only complicated cases are attended to; therefore, providers can spend more time with clients.

Our findings on the barriers to USS task-sharing with midwives resonate with several identified in ultrasound literature, including lack of training, negative attitudes among health workers, poor previous scanning experience, inadequacies in equipment, lack of maintenance for malfunctioning equipment, insufficient skilled personnel, long waiting times, long distances to service providers, equipment costs, inadequate skills of USS operators, and increased workloads [10,15–17]. The only barrier that did not emerge in our results is that of the long distances to service providers and unacceptable previous scanning experiences. Meanwhile, unmet expectations surfaced as an additional negative perspective. Our study showed that end-users [pregnant women] most often highlighted unmet expectations related to sex determination. There is a common expectation that a scan will reveal all the features of the fetus, with the sex being the most anticipated; unfortunately, this was beyond the scope of midwives which posed confusion for many pregnant women anticipating learning the sex of their babies. This aligns with findings indicating that many parents are curious whether their baby is a boy or a girl [18–20]. Additionally, our findings suggest the need for caution regarding sex determination, as it may lead to marital conflicts in cases where there is a specific desired gender for a baby. For example, in families with only boys, there may be an anticipation for a baby girl. If scans yield results contrary to the family’s expectations, it may cause conflicts among couples [18,19]. It is, therefore, important to consistently remind women before and during USS of the capabilities and limitations of health workers regarding what information can be shared with patients to mitigate unnecessary expectations and marital disputes [18].

USS task-sharing was found to be acceptable and beneficial, with perceived impacts including enhanced continuity and quality of care, improved ANC indicators, reduced complications, better maternal and neonatal outcomes, timely identification and referral of complications, confirmation of pregnancy, cost savings, and improved access. A notable benefit was reduced congestion at referral facilities, as primary-level USS reduced unnecessary referrals for basic scans[20]. This allowed referral centers to focus on complicated cases, enabling providers to spend more time with each client.

The International Confederation of Midwives (ICM) maintains a list of essential competencies for midwives, including those related to assessment of fetal status in the context of antenatal care. Each competency statement in the list includes a description of associated knowledge, skills, and behavioral indicators recommended to achieve the competency. Depending on the context and local practice requirements, additional indicators may be useful. ICM has encouraged midwifery educators, regulators, and policy makers to supplement these indicators as needed to meet in-country practice requirements. To expand ultrasound services in Malawi, the study recommends that the Nurses and Midwives Council of Malawi formally authorize USS as part of midwifery practice, and create local indicators to guide introduction of this competency into midwives’ scope of practice. Incorporating USS skills into pre-service curricula for midwives and nurses, as well as providing in-service training focused on limited obstetric USS, are essential. The scope of USS practice for midwives should be clearly defined and standardized, as recommended by ICM and in previous studies.

The study also assessed the adequacy of limited obstetric ultrasound training for midwives providing ANC services. The initial five-day theoretical and practical training, followed by mentorship and competency assessments, was recommended to be expanded by some stakeholders, who advised extending training to two weeks and developing a curriculum specifically for midwives. This aligns with other studies advocating for standardized, regulatory body-approved training[1,5–7].

Strengths of this study include its novel contribution to the literature regarding acceptability and feasibility of USS task-sharing in ANC, including a POCUS device, using robust focus group discussion (FGD) methodology, and its inclusion of diverse participant groups. Limitations include reliance on FGDs without in-depth interviews, which may have limited insights from pregnant women; the virtual format of the policy maker FGD, which introduced distractions and connectivity issues; and the lack of demographic data for individual participants, which limited subgroup analyses. Future research should consider mixed methods approaches and strategies to mitigate the challenges of virtual data collection.

## Conclusion

This study captured the perspectives and experiences of various stakeholders about task-sharing limited obstetric USS to midwives in primary health care facilities. The findings indicate that midwives, who deliver the majority of ANC in Malawi, can effectively deliver this service, with the support and endorsement of diverse stakeholders. As Malawi and other low-middle-income countries in the region plan for scale-up of this essential maternal health service, task-sharing to midwives is a viable strategy for filling human resource gaps. Community sensitization, male involvement, training for all midwives in health centers, incorporating limited obstetric USS skills into the standard midwife curriculum, and clearly defining the scope of practice for nurses will be essential.

## Data Availability

Data available on reasonable request

## Acknowledgements

We would like to acknowledge all study participants, study staff, and the Malawi Ministry of Health through the Reproductive Health Directorate, the Blantyre District Health Office, and the Queen Elizabeth Central Hospital Department of Obstetrics and Gynaecology, for their support in implementing this study.

